# Ivermectin as a potential COVID-19 treatment from the pharmacokinetic point of view: antiviral levels are not likely attainable with known dosing regimens

**DOI:** 10.1101/2020.04.11.20061804

**Authors:** Georgi Momekov, Denitsa Momekova

## Abstract

The broad-spectrum antiparasitic agent ivermectin has been very recently found to inhibit SARS-CoV-2 *in vitro* and proposed as a candidate for drug repurposing in COVID-19. In the present report the *in vitro* antiviral activity end-points are analyzed from the pharmacokinetic perspective. The available pharmacokinetic data from clinically relevant and excessive dosing studies indicate that the SARS-CoV-2 inhibitory concentrations are not likely to be attainable in humans.

## Introduction

The COVID-19 pandemics has fuelled much research efforts towards repurposing of existing drugs as possible antiviral agents, whereby the therapeutic strategies have been largely based on preexisting data for the preceding coronaviral outbreaks SARS and MERS^1-3^. The drug regulatory agencies, health authorities, key opinion leaders and policy decision makers have been significantly strained by the dilemma of evidence-based medicine and good clinical practice versus the prompt need for safe and effective treatment^4^. Unfortunately we have been witnessing huge public and political pressure for legitimation of drug-repurposing and off-label use worldwide, which nonetheless could be regarded as an acceptable compromise, pending the emergency of the current situation, but only in case of drugs with well-defined safety profiles and at least some clinical evidence in COVID-19^4,5^. Conversely most of the treatment protocols are based on observational studies and anecdotic reports^4,6-9^, albeit with a hope that the promptly emerging data from randomized studies will enable switching COVID-19 treatment back to the avenues of evidence-based medicine^10^. An exceptionally alarming phenomenon however is the public communication of drugs with preliminary *in vitro* activities against SARS-CoV-2 as potential therapeutics for COVID-19 eventually causing malignant reverberation in social media. Such example is the otherwise very interesting study of Caly et al., recently published in Antiviral Research^11^.

This paper describes the *in vitro* antiviral activity of the antiparasitic agent ivermectin in a model of Vero/hSLAM cells infected with a SARS-CoV-2 isolate (Australia/VIC01/2020)^11^. The authors have performed a pilot experiment using continuous exposure of the cells to ivermectin at 5 μmol/L and found time-dependent decrease of cell associated and supernatant viral RNA. Thereafter the antiviral activity was assessed following continuous exposure to serial dilutions of ivermectin, which caused concentration-dependent antiviral effects with practically total eradication at 5 μmol/L and half-maximal inhibition at approximately 2.5 μmol/L^11^.

The academic, virological and pharmacological impact of the newly discovered antiviral effects of ivermectin against SARS-CoV-2 is beyond any doubt. Nevertheless the notion for possible clinical translation and repurposing, which has generated enormous media coverage, needs to be carefully addressed with reference to the pharmacokinetics of ivermectin. In this paper we sought to analyze the dosing regimens of the drug, the available maximal plasma concentration levels to allow detailed juxtaposition with the SARS-CoV-2 inhibitory effects and to question the paradigm for the plausibility of ivermectin repurposing in COVID-19.

## Materials and methods

A literature survey was performed in order to analyze the published dose regimens and to collect human exposure data for ivermectin, following clinically relevant (150 – 800 μg/kg) or excessive dosing (up to 2000 μg/kg). The available pharmacokinetic data for ivermectin in patients with parasitic infection and healthy volunteers were pooled and the maximal plasma concentration levels (C_max_) used as surrogates for juxtaposition with the *in vitro* SARS-CoV-2 inhibitory findings. The published concentrations showing antiviral activity were recalculated in ng/ml to allow direct comparison with the pharmacokinetic data.

## Results and discussion

Ivermectin has a valuable clinical role for the management of different parasitic diseases whereby the described therapeutic regimens, could be summarized as follows: 150 μg/kg once yearly for treatment of onchocerciasis, 200 μg/kg as a single dose for strongyloidiasis, 150 to 200 μg/kg twice yearly or alternatively 300 to 400 μg/kg once yearly in endemic areas for lymphatic filariasis, and 200 μg/kg in conjunction with topical drugs for hyperkeratotic, also known as crusted or ‘Norwegian’ scabies^12-14^.

Ivermectin is a semisynthetic analogue of the natural product avermectin B_1a_, a lipophilic macrolide isolated from *Streptomyces avermitilis* developed as a crop management insecticide. Ivermectin affects a plethora of ivertebrate species, incl. parasitic nematodes, arachnids and insects. Its mode of action on target species is by potentiating GABA-mediated neurotransmission and by binding to glutamate-gated Cl^-^ channels, found only in invertebrates^13^. The drug induces tonic paralysis of the musculature of susceptible parasites, and eventually death^13^. At the recommended doses, ivermectin does not readily penetrate the central nervous system (CNS) of mammals, where GABA functions as a neurotransmitter^13,15^. Conversely, in healthy volunteers and infected patients, the drug is usually well tolerated at the therapeutic dose ranges^12-14^. A recent meta-analysis has shown that even larger doses (up to 800 μg/kg) with a several years period of follow-up could be well tolerated in patients with parasitic infections^16^. The largest dose intensity with registered pharmacokinetic parameters in healthy subjects is 120 mg, corresponding to up to 2000 μg/kg^12^.

As evident from the analyzed pharmacokinetic data, both the clinically applied dosage schedules and the aforementioned excessive 120 mg dose yield blood levels at the ng/mL i.e. nanomolar range (Table 1). These concentrations are orders of magnitude lower, as compared to the *in vitro* antiviral end-points described in the study of Caly et al^11^. Table 2 summarizes the *in vitro* inhibitory concentrations recalculated in ng/mL (based on a molecular weight of 875.1) to allow direct juxtaposition with the pharmacokinetic parameters in Table 1. Moreover the *in vitro* data were compared to the C_max_ values obtained after 36 mg and 120 mg doses corresponding to dose intensities of up to 700 μg/kg^17^ or 2000 μg/kg^12^ respectively, with calculation of the corresponding exposure ratios.

**Table 1.** Published pharmacokinetic parameters for ivermectin following clinically relevant and excessive dosing

| Pharmacokinetic study | Dose | Absorption parameters |  |  |  | Elimination |  |
| --- | --- | --- | --- | --- | --- | --- | --- |
| | | $C_{\max}$<br>(ng/mL) | $C_{\max}$<br>(nmol/L) | $t_{\max}$ (h) | $t_{1/2(\text{abs})}$ (h) | $t_{1/2}$ (h) | Cl<br>(1 kg<sup>-1</sup>·day<sup>-1</sup>) |
| Krishna et al., 1993<sup>21</sup> | 6 mg (tablets) | 23.1 | 26.4 | 4.3 | 0.5 | 12.6 | 4.28 |
| Krishna et al., 1993<sup>21</sup> | 12 mg (tablets) | 30.4 | 34.7 | 10.3 | 2.5 | 13.4 | 4.03 |
| Long et al., 2001<sup>22</sup> | 6 mg (tablets) | 20.2 | 23.1 | 4.7 | 1.4 | 11.1 | 7.57 |
| Long et al., 2001<sup>22</sup> | 12 mg (tablets) | 23.5 | 26.9 | 5.3 | 1.4 | 21.1 | 6.53 |
| Long et al., 2001<sup>22</sup> | 18 mg (tablets) | 31.2 | 35.7 | 5.1 | 1.7 | 16.7 | 10.6 |
| Edwards et al., 1988<sup>23</sup> | 12 mg (solution) | 81 | 92.6 | 3.6 | – | – | – |
| Edwards et al., 1988<sup>23</sup> | 12 mg (tablets) | 50 | 57.1 | 3.4 | – | – | – |
| Edwards et al., 1988<sup>23</sup> | 12 mg (capsules) | 46 | 52.6 | 3.6 | – | – | – |
| Ogbuokiri et al., 1993<sup>24</sup> | 150 µg/kg | 37.9 | 43.3 | – | – | – | – |
| Baraka et al., 1996<sup>25</sup> | 150 µg/kg | 52.2 | 59.7 | 5.2 | – | 35.0 | – |
| Elkassaby, 1991<sup>26</sup> | 150 µg/kg | 39 | 44.6 | 5.6 | – | 16 | – |
| Okonkwo, et al., 1993<sup>27</sup> | 6 mg (tablets) | 38.2 | 43.7 | 4.7 | – | 54.5 | 3.1 |
| Njoo, et al. 1995<sup>28</sup> | 150 µg/kg | - | - | - | - | 19.9 | – |
| Muñoz, et al., 2018<sup>17</sup> | 36 mg (550 - 700 µg/kg) | 96.2 | 109.9 | 3.64 | - | 91.77 |  |
| Guzzo et al., 2002<sup>12</sup> | 90 mg | 158.1 | 180.7 | 4.9 | - | 18.8 | - |
| Guzzo et al., 2002<sup>12</sup> | 120 mg (1404 - 2000 µg/kg) | 247.8 | 283.2 | 4.2 | - | 19.1 | - |

**Table 2.** Comparison between the in vitro inhibitory concentration of ivermectin^11^ with exemplar published C_max_ values of the drug^12,17^

| *In vitro* end-points<sup>11</sup> | Inhibitory concentrations (µmol/L) | Inhibitory concentrations recalculated in µg/mL | Inhibitory concentrations recalculated in ng/mL | Ratio between the inhibitory concentrations and clinically relevant C<sub>max</sub> (96.2 ng/mL) | Ratio between the inhibitory concentrations and C<sub>max</sub> after excessive dosing 247.8 (ng/mL) |
| --- | --- | --- | --- | --- | --- |
| IC<sub>50</sub> - cell associated (E gene)* | 2.8 | 2.45 | 2450 | 25.5 | 9.9 |
| IC<sub>50</sub> - supernatant (E gene) | 2.4 | 2.1 | 2100 | 25.9 | 8.5 |
| IC<sub>50</sub> -cell associated (RdRp gene)** | 2.5 | 2.19 | 2190 | 27.0 | 8.8 |
| IC<sub>50</sub> - supernatant (RdRp gene) | 2.2 | 1.95 | 1950 | 24.1 | 7.9 |
| Average IC<sub>50</sub> | 2.475 | 2.166 | 2166 | **22.51** | **8.74** |
| IC<sub>90-100</sub> (highest and most effective concentration tested) | 5.0 | 4.3755 | 4375 | **55.0** | **17.7** |

\*E - envelope protein gene; \*\* RdRp - RNA-dependent-RNA-polymerase gene.<sup>11</sup>

The analyzed data show that, at least at the clinically relevant dose ranges of ivermectin, the published *in vitro* inhibitory concentrations and especially the 5 μmol/L level causing almost total disappearance of viral RNA are virtually not achievable with the heretofore known dosing regimens in humans. The 5 μmol/L concentration is over 50 times higher than the levels attainable after 700 μg/kg^17^ and 17 times higher vs. the largest C_max_ found in the literature survey (247.8 ng/ml)^12^. Moreover the authors’ claim for achieving viral inhibition with a single dose is inappropriate because practically the infected cells have been continuously exposed at concentrations that are virtually unattainable even with excessive dosing of the drug. In other words, the experimental design is based on clinically irrelevant drug levels with inhibitory concentrations whose targeting in a clinical trial seems doubtful at best.

The mechanistic rationale could be questioned as well – ivermectin has been proposed to mediate its antiviral effects via host cells mechanisms, namely inhibition of the importin α/β-mediated nuclear transfer of viral cargo proteins^18-20^. The clinical translation of this mechanism however is greatly debatable as for instance the IC_50_ for importin α/β binding to a DENV non-structural protein is 17 μmol/L^18^.

The repurposed antimalarial drugs hydroxychloroquine and chloroquine that have been included in numerous COVID-19 treatment protocols also have micromolar inhibitory concentrations against SARS-CoV-2^29^ and nanomolar C_max_ values^30^. Nevertheless these agents have enormous apparent volumes of distribution and presumably disproportionally larger tissue levels relative to the plasma concentrations, which makes the translation of the *in vitro* data plausible^30-32^. The other very promising agent tested in clinical trials and applied as compassionate use for COVID-19 is remdesivir^1,33^. This broad-spectrum antiviral drug originally developed for Ebola shows potent inhibitory effects against SARS-CoV-2 with an IC_50_ of 0.77 μmol/L (464 ng/mL)^34^. This concentration is readily attainable as the drug is given as venous infusion. The typical dosing schedule for remdesivir – initial infusion of 200 mg, followed by 100 mg/daily for a total of 5 days – yields maximal plasma concentrations of 5440 ng/mL in the first day and 2610 on day 5 ^33^.

In the case of ivermectin, however, the potential repurposing plausibility if any is at present not very likely, because the antiviral concentrations would be attainable only after massive overdose. The therapeutic application of ivermectin is usually not associated with significant toxicity, whereby the majority of documented adverse effects, such as: nausea, rash, dizziness, itching, eosinophilia, abdominal pain, fever, tachycardia, could be generally attributable to the gross lethality of invading microfilarias giving rise to Mazzotti-like reactions^13,14,35^. Nevertheless, at large doses, the drug could penetrate the blood-brain barrier and could affect GABA-ergic transmission causing CNS depression and potentiation of the effects of benzodiazepines^15^. Human exposures at doses multifold higher than the therapeutic one is expected to give rise to side effects similar to those documented in preclinical mammalian testing^13,15^, noteworthy the anti-SARS-CoV-2 levels overlap with the inhibitory concentrations of ivermectin on P-glycoprotein, and other ATP-binding cassette transporters^36^, which limit the penetration of the drug in the CNS^15,37^. Human overdoses have been associated with vomiting, tachycardia and ECG abnormalities, significant blood pressure fluctuations, CNS effects (drowsiness, ataxia) and visual disturbances (mydriasis). Accidental self-injection of a veterinary medicinal product has produced signs of clinical toxicity, albeit the drug was applied at therapeutically relevant dose (approximately 200 μg/kg)^15^.

It has to be emphasized that general public communication of drugs as potential COVID-19 therapeutics, based solely on *in vitro* data, is neither scientifically nor ethically appropriate. Ivermectin has been previously shown to exert antiviral activity *in vitro* against Dengue fever virus (DENV)^20^, influenza virus, West Nile Virus, Venezuelan equine encephalitis virus, and heralded as a possible antiviral drug, but so far there has not been any clinical translation of these data. Noteworthy, a clinical trial for the treatment of Dengue fever in Thailand failed to show clinical benefits^11^. In light of the aforementioned pharmacokinetic considerations, this is not surprising given that the published inhibitory concentrations against DENV1-4 ranged within 1.66 - 2.32 μmol/L^20^.

The presented data analysis is an oversimplification of the pharmacokinetic processes because it is based solely on total C_max_ as a drug exposure surrogate. To address this issue, a Supplemental data file has been compiled to analyze the influence of the tissue accumulation and protein binding of ivermectin.

The world has already seen epidemics of self-medication, drug shortages and even overdoses with chloroquine and hydroxychloroquine^39,40^. Unfortunately the Caly et al. study, which prompted enormous public interest, has the potential to evoke similar tragic sequels, especially having in mind that in many countries the drug is only available as solutions for injection for veterinary use, whose potential for serious toxicological outcomes in humans is undisputed. In Bulgaria, in particular this study has prompted the National Veterinarians’ Union to share their concerns about the hysteria this study has evoked and to firmly discourage self-medication with ivermectin, which in this country is only available for use in animals^41^. In a 10^th^ April letter to stakeholders FDA has similarly shared its concerns on this issue and explicitly advised against any attempts for self-medication with ivermectin for COVID-19^42^.

Similar general pharmacological considerations and toxicological concerns have been shared in other recent contributions, questioning the rationale for ivermectin repurposing for SARS-CoV-2 infection^43,44^. Nevertheless, at the time of the writing of this paper several clinical trials of ivermectin for COVID-19 have been registered^45^ and an international registry-based case-control study has been described in a non-peer reviewed preprint^46^.

## Conclusions

The available pharmacokinetic data for ivermectin indicate that at the doses routinely used for the management of parasitic diseases the SARS-CoV-2 inhibitory concentrations are practically not attainable. At present any empiric treatment with ivermectin or its inclusion in therapeutic protocols are not scientifically justifiable. The very consideration of the drug as a broad spectrum antiviral agent is incorrect because it has failed to demonstrate antiviral effects beyond the *in vitro* level. Pending the paucity of reliable data from controlled studies and the aforementioned pharmacokinetic considerations the application of ivermectin in COVID-19 patients is to be decisively discouraged.

## Data Availability

The manuscript is based on survey of pharmacokinetic and virological data available in the published literature.

## References

1. Li G, De Clercq E. Therapeutic options for the 2019 novel coronavirus (2019-nCoV). Nat Rev Drug Discov 2020;19:149–150.

2. De Clercq E. Potential antivirals and antiviral strategies against SARS coronavirus infections. Expert Rev Anti Infect Ther 2006;4:291–302.

3. Shereen MA, Khan S, Kazmi A, Bashir N, Siddique R. COVID-19 infection: Origin, transmission, and characteristics of human coronaviruses. Journal of Advanced Research 2020;24:91–98.

4. Interim clinical guidance for patients suspected of/confirmed with COVID-19 in Belgium - 31 march 2020; version 6. 2020. at https://epidemio.wiv-isp.be/ID/Documents/Covid19/COVID-19InterimGuidelinesTreatmentENG.pdf.) [cited 2020 Apr 16]

5. Therapeutics and clinical trials. In: Brigham & Women’s Hospital COVID-19 Clinical Guidelines. 2020. at https://covidprotocols.org/protocols/04-therapeutics-and-clinical-trials#systemic-corticosteroids.) [cited 2020 Apr 16]

6. Tratamientos disponibles para el manejo de la infección respiratoria por SARS-CoV-2 (fecha de actualización: 28 de marzo de 2020). Agencia Española de Medicamentos y Productos Sanitarios (AEMPS) 2020: https://www.aemps.gob.es/la-aemps/ultima-informacion-de-la-aemps-acerca-del-covid%E2%80%9119/tratamientos-disponibles-para-el-manejo-de-la-infeccion-respiratoria-por-sars-cov-9112/?lang=en [cited 2020 Apr 9116].

7. Medicamenteuze behandelopties bij patiënten met COVID-19 (infecties met SARS-CoV-2). Stichting Werkgroep Antibiotica Beleid (SWAB) 2020: https://swab.nl/nl/article/nieuws/494/voorlopige-behandelopties-covid-419-infecties-met-sars-cov-492 [cited 2020 Apr 2016].

8. Vademecum per la cura delle persone con malattia da COVI-19. Società Italiana di Malattie Infettive e Tropicali 2020: http://www.simit.org/medias/1569-covid1519-vademecum-1513-1503-1202.pdf [cited 2020 Apr 1516].

9. Liang T, ed. Handbook of COVID-19 Prevention and Treatment Zhejiang First Affiliated Hospital, Zhejiang University School of Medicine; 2020.

10. Thailand joins the WHO “Solidarity Trial”: global testing of effective treatments of COVID-19 across 8 countries—an aggressive effort to save lives from the pandemic. World Health Organization 2020: https://www.who.int/thailand/news/detail/20-03-2020-thailand-joins-the-who-solidarity-trial-global-testing-of-effective-treatments-of-covid-2019-across-2028-countries-an-aggressive-effort-to-save-lives-from-the-pandemic [cited 2020 Apr 2016].

11. Caly L, Druce JD, Catton MG, Jans DA, Wagstaff KM. The FDA-approved Drug Ivermectin inhibits the replication of SARS-CoV-2 in vitro. Antiviral Res 2020 [cited 2020 Apr 16]:104787. DOI: 104710.101016/j.antiviral.102020.104787.

12. Guzzo CA, Furtek CI, Porras AG, et al. Safety, tolerability, and pharmacokinetics of escalating high doses of ivermectin in healthy adult subjects. J Clin Pharmacol 2002;42:1122–1133.

13. Loukas A, Hotez PJ. Chemotherapy of helminth infections. In: Brunton LL, Lazo JS, Parker KL, eds. Goodman & Gilman’s The Pharmacological Basis of Therapeutics, 11^th^ Ed. New York: McGraw Hill; 2006: 1073-1093.

14. British National Formulary 60. London: BMJ Group/Pharmaceutical Press; 2010.

15. Ivermectin (INCHEM Monograph). 1992. at http://www.inchem.org/documents/pims/pharm/ivermect.htm.) [cited 2020 Apr 16]

16. Navarro M, Camprubí D, Requena-Méndez A, et al. Safety of high-dose ivermectin: a systematic review and meta-analysis. J Antimicrob Chemother 2020;75:827–834.

17. Muñoz J, Ballester MR, Antonijoan RM, et al. Safety and pharmacokinetic profile of fixed-dose ivermectin with an innovative 18mg tablet in healthy adult volunteers. PLOS Neglected Tropical Diseases 2018 [cited 2020 Apr 16];12:e0006020. DOI: 0006010.0001371/journal.pntd.0006020. eCollection 0002018 Jan.

18. Yang SNY, Atkinson SC, Wang C, et al. The broad spectrum antiviral ivermectin targets the host nuclear transport importin alpha/beta1 heterodimer. Antiviral Res 2020 [cited 2020 Apr 16];177:104760. DOI: 104710.101016/j.antiviral.102020.104760.

19. Lundberg L, Pinkham C, Baer A, et al. Nuclear import and export inhibitors alter capsid protein distribution in mammalian cells and reduce Venezuelan Equine Encephalitis Virus replication. Antiviral Res 2013;100:662–672.

20. Tay MYF, Fraser JE, Chan WKK, et al. Nuclear localization of dengue virus (DENV) 1–4 non-structural protein 5; protection against all 4 DENV serotypes by the inhibitor Ivermectin. Antiviral Research 2013;99:301–306.

21. Krishna DR, Klotz U. Determination of ivermectin in human plasma by high-performance liquid chromatography. Arzneimittelforschung 1993;43:609–611.

22. Long QC, Ren B, Li SX, Zeng GX. Human pharmacokinetics of orally taken ivermectin. Chin J Clin Pharmacol 2001;17:203–206.

23. Edwards G, Dingsdale A, Helsby N, Orme ML, Breckenridge AM. The relative systemic availability of ivermectin after administration as capsule, tablet, and oral solution. Eur J Clin Pharmacol 1988;35:681–684.

24. Ogbuokiri JE, Ozumba BC, Okonkwo PO. Ivermectin levels in human breastmilk. Eur J Clin Pharmacol 1993;45:389–390.

25. Baraka OZ, Mahmoud BM, Marschke CK, Geary TG, Homeida MM, Williams JF. Ivermectin distribution in the plasma and tissues of patients infected with Onchocerca volvulus. Eur J Clin Pharmacol 1996;50:407–410.

26. Elkassaby MH. Ivermectin uptake and distribution in the plasma and tissue of Sudanese and Mexican patients infected with Onchocerca volvulus. Trop Med Parasitol 1991;42:79–81.

27. Okonkwo PO, Ogbuokiri JE, Ofoegbu E, Klotz U. Protein binding and ivermectin estimations in patients with onchocerciasis. Clin Pharmacol Ther 1993;53:426–430.

28. Njoo FL, Beek WM, Keukens HJ, et al. Ivermectin detection in serum of onchocerciasis patients: relationship to adverse reactions. Am J Trop Med Hyg 1995;52:94–97.

29. Colson P, Rolain JM, Lagier JC, Brouqui P, Raoult D. Chloroquine and hydroxychloroquine as available weapons to fight COVID-19. Int J Antimicrob Agents 2020 [cited 2020 Apr 16];55:105932. DOI: 105910.101016/j.ijantimicag.102020.105932.

30. Thummel KE, Shen DD, Isoherannen N, Smith HE. Appendix II. Design and optimization of dosage regimens. In: Brunton LL, Lazo JS, Parker KL, eds. Goodman & Gilman’s The Pharmacological Basis of Therapeutics, 11^th^ Ed. New York: McGraw Hill; 2006: 1787-1888.

31. Shapiro TA, Goldberg DE. Chemotherapy of protozoal infections: malaria. In: Brunton LL, Lazo JS, Parker KL, eds. Goodman & Gilman’s The Pharmacological Basis of Therapeutics, 11^th^ Ed. New York: McGraw Hill; 2006: 1021-1047.

32. Yao X, Ye F, Zhang M, et al. In vitro antiviral activity and projection of optimized dosing design of hydroxychloroquine for the treatment of severe acute respiratory syndrome coronavirus 2 (SARS-CoV-2). Clinical Infectious Diseases 2020 [cited 2020 Apr 16];ciAA237:DOI: 10.1093/cid/ciaa1237.

33. European Medicines Agency /178637/2020/Human Medicines Division. Summary on compassionate use - Remdesivir Gilead, International Nonproprietary Name: remdesivir, Procedure No. EMEA/H/K/5622/CU (03 Apr. 2020). 2020. at https://www.ema.europa.eu/en/documents/other/summary-compassionate-use-remdesivir-gilead_en.pdf.) [cited 2020 Apr 16]

34. Wang M, Cao R, Zhang L, et al. Remdesivir and chloroquine effectively inhibit the recently emerged novel coronavirus (2019-nCoV) in vitro. Cell Res 2020;30:269–271.

35. Fawcett RS. Ivermectin use in scabies. Am Fam Physician 2003;68:1089–1092.

36. Lespine A, Dupuy J, Orlowski S, et al. Interaction of ivermectin with multidrug resistance proteins (MRP1, 2 and 3). Chemico-biological interactions 2006;159:169–179.

37. Chaccour C, Hammann F, Ramón-García S, Rabinovich NR. Ivermectin and Novel Coronavirus Disease (COVID-19): Keeping Rigor in Times of Urgency. The American Journal of Tropical Medicine and Hygiene 2020.

38. Götz V, Magar L, Dornfeld D, et al. Influenza A viruses escape from MxA restriction at the expense of efficient nuclear vRNP import. Scientific Reports 2016 [cited 2020 Apr 16];6:23138. DOI: 23110.21038/srep23138.

39. Nigeria records chloroquine poisoning after Trump endorses it for coronavirus treatment. 2020 at https://edition.cnn.com/2020/03/23/africa/chloroquine-trump-nigeria-intl/index.html.) [cited 2020 Apr 16]

40. Jakhar D, Kaur I. Potential of chloroquine and hydroxychloroquine to treat COVID-19 causes fears of shortages among people with systemic lupus erythematosus. Nature Medicine 2020 [cited 2020 Apr 16];26:632. DOI:610.1038/s41591-41020-40853-41590.

41. Veterinarians warn: do not buy ivermectin for treatment of humans, it is very toxic. 2020. at https://news.bnt.bg/news/veterinari-preduprezhdavat-ne-kupuvaite-ivermektin-za-lechenie-na-hora-silno-toksichen-e-1047797news.html.) [cited 2020 Apr 16]

42. FDA letter to stakeholders: Do not use ivermectin intended for animals as treatment for COVID-19 in humans (April 10, 2020). 2020. at https://www.fda.gov/animal-veterinary/product-safety-information/fda-letter-stakeholders-do-not-use-ivermectin-intended-animals-treatment-covid-19-humans.) [cited 2020 Apr 16]

43. Bray M, Rayner C, Noël F, Jans D, Wagstaff K. Ivermectin and COVID-19: a report in Antiviral Research, widespread interest, an FDA warning, two letters to the editor and the authors’ responses. Antiviral Res 2020 [cited 2020 May 21]:104805. DOI:104810.101016/j.antiviral.102020.104805.

44. Schmith VD, Zhou JJ, Lohmer LR. The approved dose of ivermectin alone is not the ideal dose for the treatment of COVID-19. Clin Pharmacol Ther 2020 [cited 2020 Apr 16]:DOI: 10.1002/cpt.1889.

45. U.S. National Library of Medicine: ClinicalTrials.gov. 2020. at https://clinicaltrials.gov/ct2/results?cond=covid-19&term=ivermectin&cntry=&state=&city=&dist=) [cited 2020 May 21]

46. Usefulness of Ivermectin in COVID-19 Illness (April 19, 2020). 2020. at https://ssrn.com/abstract=3580524.) [cited 2020 May 21]

